# Digital biomechanical biomarkers in the clinical assessment of patients with peripheral neuropathies

**DOI:** 10.1101/2024.10.15.24315365

**Authors:** Clara Tejada-Illa, Jordi Pegueroles, Mireia Claramunt-Molet, Ariadna Pi-Cervera, Ainhoa Heras-Delgado, Jesus Gascón-Fontal, Sebastian Idelsohn-Zielonka, Mari Rico, Nuria Vidal, Lorena Martín-Aguilar, Marta Caballero-Ávila, Cinta Lleixà, Roger Collet-Vidiella, Laura Llansó, Álvaro Carbayo, Ana Vesperinas, Luis Querol, Elba Pascual-Goñi

**Author notes:** **These authors contributed equally to this work.** (Clara Tejada-Illa, Jordi Pegueroles). Correspondence to: Luis Querol Neuromuscular Diseases Unit - Department of Neurology Hospital de la Santa Creu i St. Pau Sant Quintí 77 (2nd Floor) 08041 Barcelona.

## Abstract

The clinical status and treatment response of patients with peripheral neuropathies (PN) rely on subjective and inaccurate clinical scales. Wearable sensors have shown success in evaluating gait and balance in individuals with other neurological disorders. We aimed to explore the ability of biomechanical analysis via wearable technology to monitor disease activity in PN by conducting a single-center, longitudinal study to analyze gait parameters in PN patients and healthy controls using wearable sensors. First, we analyzed the sensor’s ability to detect changes in ataxia and steppage gait severity and found significant differences in spatiotemporal and angular variables. Second, we found correlations between biomechanical features and clinical scales linked to specific gait phenotypes. Finally, we demonstrated that this technology captures clinical changes in gait features over time. Our study provides proof-of-concept that wearable technology effectively detects and grades gait impairment, captures clinically relevant changes, and could enhance gait assessment in routine care and research for PN patients.

## Introduction

Peripheral neuropathies (PN) are a heterogeneous group of diseases of the peripheral nervous system^1–3^ that present with diverse symptoms, including weakness, sensory disturbances, ataxia, fatigue, and pain, that frequently cause gait dysfunction^4–7^. These diseases lack objective prognostic and disease activity biomarkers^8–10;^ assessments of clinical status and disease activity are based on clinical scales, which are often imprecise^11,12^, with significant ceiling and floor effects and fluctuations in stable patients^13^, which are associated with substantial placebo effects^14^, and fail to capture minor disease changes that could reflect ongoing nerve damage. In the long term, this imprecision results in inefficient care and, eventually, irreversible disability^15^. Thus, there is an unmet need in the field to develop objective outcome measures to quantify and monitor disease parameters^9,16,17^.

In recent years, new technologies, such as portable inertial sensors that are able to measure motor capacity^18,19^ and gait, have been developed^20,21^. These sensors are placed on the patient’s body and are incorporated into insoles, bracelets, or clothing^22,23^, which allows the unbiased measurement of diverse biomechanical parameters while performing motor tasks^24,25^. Wearable sensors may provide greater objectivity^26^ and measurability^27^ in the assessment of the clinical status of patients with neurological conditions^28–31^, helping overcome the limitations of current monitoring strategies^32,33^. Moreover, the development of affordable technologies capable of automatically recognizing and monitoring disease status may improve disease care and reduce the load on the healthcare system^29,34,35^.

Multiparametric wearable technologies have not been tested in patients with PN. Our main objective was to test the ability of a set of digital biomechanical biomarkers (DBB), consisting of multiple spatiotemporal features, biomechanical angles, and plantar pressure parameters, to monitor the clinical status of patients with PN. To achieve this goal, three objectives were developed. First, we investigated whether DBB could detect differences from healthy controls in two frequent gait phenotypes that appear in patients with PNs: gait ataxia and steppage gait. Second, we analyzed whether DBB was correlated with objective and subjective clinical scales. Third, we tested the ability of DBB to capture clinically significant changes over time.

## Methods

### Patients

Patients with PN followed in the Neuromuscular Diseases Unit of our center were included in the study. The mean age was 62.2 years, and 51 patients (60.7%) were male. We recruited 37 chronic inflammatory demyelinating polyneuropathy (CIDP) patients (3.5%), 3 chronic ataxic neuropathy, ophthalmoplegia, immunoglobulin M [IgM] paraprotein (CANOMAD) patients (3.5%), 21 monoclonal gammopathy patients of undetermined significance associated with IgM (IgM-MGUS) patients (25%), 7 patients with autoimmune nodopathies (8.3%), 11 patients with hereditary neuropathies (13.1%), and 50 healthy controls. Patients were classified by the evaluating neurologist according to their gait pattern into three gait groups (gait ataxia, steppage or normal gait). Additionally, the severity of ataxia and steppage (mild, moderate, or severe) was classified according to prespecified criteria (Supplementary Table 1).

This study was conducted according to a protocol approved by the Ethics Committee of the Hospital de la Santa Creu i Sant Pau (code IIBSP-NMI-2019-107). All patients provided written informed consent to participate in the study.

### Protocol of the study

Patients were monitored for two years. Visits were scheduled every six months for stable patients and every three months for patients in which a relapse or treatment change occurred and every six months again upon stabilization. The protocol scheme is summarized in Fig. 1. During the visits, the evaluating neurologists performed a thorough neurological evaluation including the following scales or scores: Medical Research Council sum score (MRCss), Inflammatory Neuropathy Cause and Treatment (INCAT), Modified Internacional Cooperative Ataxia Rating Scale and Scale for the Assessment and Rating of Ataxia (MICARS-SARA) and grip strength testing via a Martin vigorimeter. After the neurological evaluation, patients completed the Inflammatory Rasch-built Overall Disability Scale (iRODS) questionnaire. Finally, the 2-minute walking test (2MWT) was performed while the participants wore all the biomechanical sensors.

**Figure 1.**
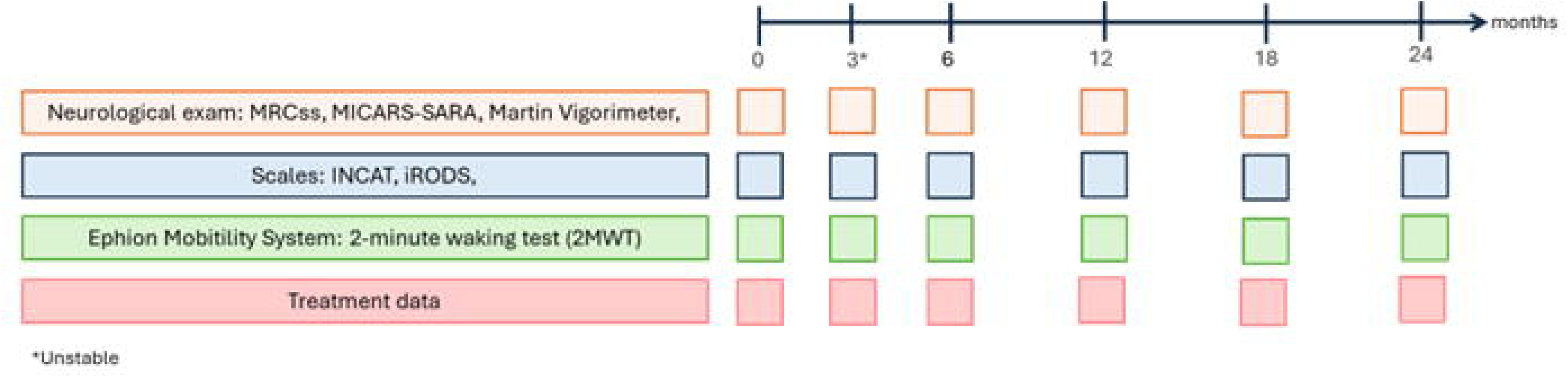
Protocol of the study. Monitoring visits performed every 6 months during 2 years for those patients who remained stable and for those who presented a relapse or a treatment change, a 3-month visit (3*) was performed after that date. MRCss: Medical Research Council sum score. INCAT: Inflammatory Neuropathy Cause and Treatment. iRODS: Inflammatory Rasch-built Overall Disability Scale. MICARS-SARA: Modified Intemacional Cooperative Ataxia Rating Scale and Scale for the Assessment and Rating of Ataxia.

### Wearable technology

We used the Ephion Mobility system^36^ (Ephion Health, Barcelona, Spain), which integrates multiple parameter sensors, including 7 inertial sensors (Movesense, Vantaa, Finland^37^) placed at different locations (ankles, thigh, and chest) and insoles with inertial and plantar pressure sensors (Moticon, Munich, Germany^38^). The full protocol included pants with integrated surface EMGs, but for the purpose of this study, these data was not analyzed. All the sensors are synchronized via WT+ software on a smartphone (Supplementary Fig. 1). This biomechanical sensor system enables the identification of different variables, such as kinematic, foot, ankle, knee, and hip angle flexion measurements; spatiotemporal variables, such as velocity, double support time, stride length and cadence; and plantar pressure variables, such as vertical force and center of pressure (COP) and heart rate. Information about which variables and features are measured with each sensor and their biomechanical interpretation are detailed in supplementary table 5. Individuals wore the wearable system while performing 2MWT. The data from the entire walking test were divided into gait cycles. A gait cycle describes a cyclical walking pattern; it starts when one foot contacts the ground and ends when the same foot contacts the ground again.

### Statistical analysis

The subjects’ gait parameters were assessed by taking the mean of all gait cycles per parameter. To assess differences between the control and pathological groups (ataxia, steppage and whole cohort), a Kruskal‒Wallis test was performed, and multiple comparisons were performed (false discovery rate - FDR), followed by post hoc comparisons via Tukey’s HSD test. A significance level of corrected p value<0.0001 was established to select features that effectively differentiated these patient groups from control individuals.

Additionally, linear mixed-effects models (LME) were used to assess the relationships of the computed gait parameters with ataxia, steppage or both gait patterns. To model the relationship between clinical impact and the different gait parameters obtained in the 2MWT, we used LME using clinical scales, iRODS and the 2MWT distance as fixed effects. A significance level of corrected p value<0.05 was established to select features that effectively showed correlations for gait patterns and for clinical scales.

Finally, the Wilcoxon test and LME were used to capture clinically significant longitudinal changes in gait parameters only in patients who experienced significant clinical changes. A significance level of corrected p value<0.0001 was established to select features that effectively differentiated these patient groups from control individuals. Patients selected for these analyses included those who presented clinically meaningful worsening or improvement, defined as (1) a 2-point or greater change in the MRC score and/or (2) a 4-point change on the iRODS scale^39^, which are widely accepted minimal clinically important changes in patients with PN. For Wilcoxon analysis, the best and worst clinical assessments were selected to study the longitudinal clinical changes. For all LME analyses, participant-specific intercepts and slopes were added as random factors, and the FDR was also used for multiple comparisons correction. The p values and magnitude of each test are represented in the figures and not in the text for readability reasons.

## Results

### Baseline clinical characteristics

For the analysis of differences in normal and abnormal gait patterns, we included 48 patients with ataxia (40 mild, 6 moderate and 2 severe), 34 with steppage (23 mild, 5 moderate and 6 severe) and 50 controls who underwent the 2MWT. Among these patients, 30 presented mixed gait patterns with ataxia plus steppage. Additionally, patients who presented normal gait patterns were excluded from this first analysis. The number of patients in each group is summarized in supplementary Table 2. The mean age of the patients included in the ataxia group was 57.4 years and 33 patients (68.8%) were male. The mean age of the patients included in the steppage group was 59.3 years and 20 patients (58.8%) were male. The baseline results of the 2MWT, vigorimeter grip strength, MRCss, INCAT and iRODS scores of the included patients are summarized in Table 1 and supplementary Table 3 for the ataxia group and in supplementary Table 4 for the steppage group. For the analysis of the correlations between gait patterns and DBB and for the analysis of correlations between conventional scales and DBB, we included 79 patients with ataxia and/or steppage or normal gait patterns. For the longitudinal study, we included 31 patients who presented a change in MRCss and 22 patients who presented a change in the iRODS scale. The number of tests included in the analysis of differences between normal and abnormal gait patterns and in the longitudinal study are summarized in Supplementary Table 5.

**Table 1.**
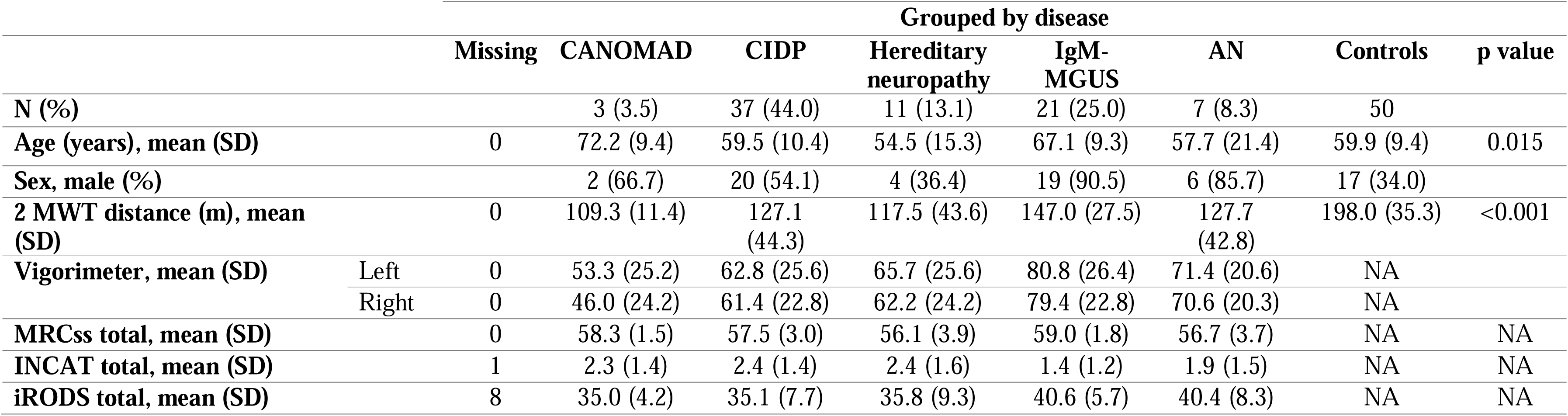
Patients’ classification according to their disease. Number, mean, standard deviation (sd) of patients included of each pathology according to age and sex. Mean and sd results of 2-minute-walking-test (2MWT), grip strength using vigorimeter, Medical Research Council sum score (MRCss), Inflammatory Neuropathy Cause and Treatment (INCAT), Inflammatory Rasch-built Overall Disability Scale (iRODS). Chi-quadrat test used for study the differences between sex and ANOVA test used to analyze the differences between controls and patients in 2MWT. CANOMAD: chronic ataxic neuropathy, ophthalmoplegia, immunoglobulin M [IgM] paraprotein, cold agglutinins, and disialosyl antibodies; CIDP: chronic inflammatory demyelinating polyneuropathy; IgM-MGUS: monoclonal gammopathy of undetermined significance associated with IgM; AN: autoimmune nodopathy.

### DBB captures differences between normal and abnormal gait patterns

The spatiotemporal features (ST) velocity, stride duration and length, cadence, and double support time were analyzed for both groups of patients (Fig. 2). In patients with ataxia (Fig. 2A), velocity, stride length and cadence decreased significantly across the different severity groups. The opposite occurred with stride duration and double support time, which increased significantly in the different severity groups. The same occurred for the group of steppage patients (Fig. 2B), in whom we observed the same trends as those in the ataxia group.

**Figure 2.**
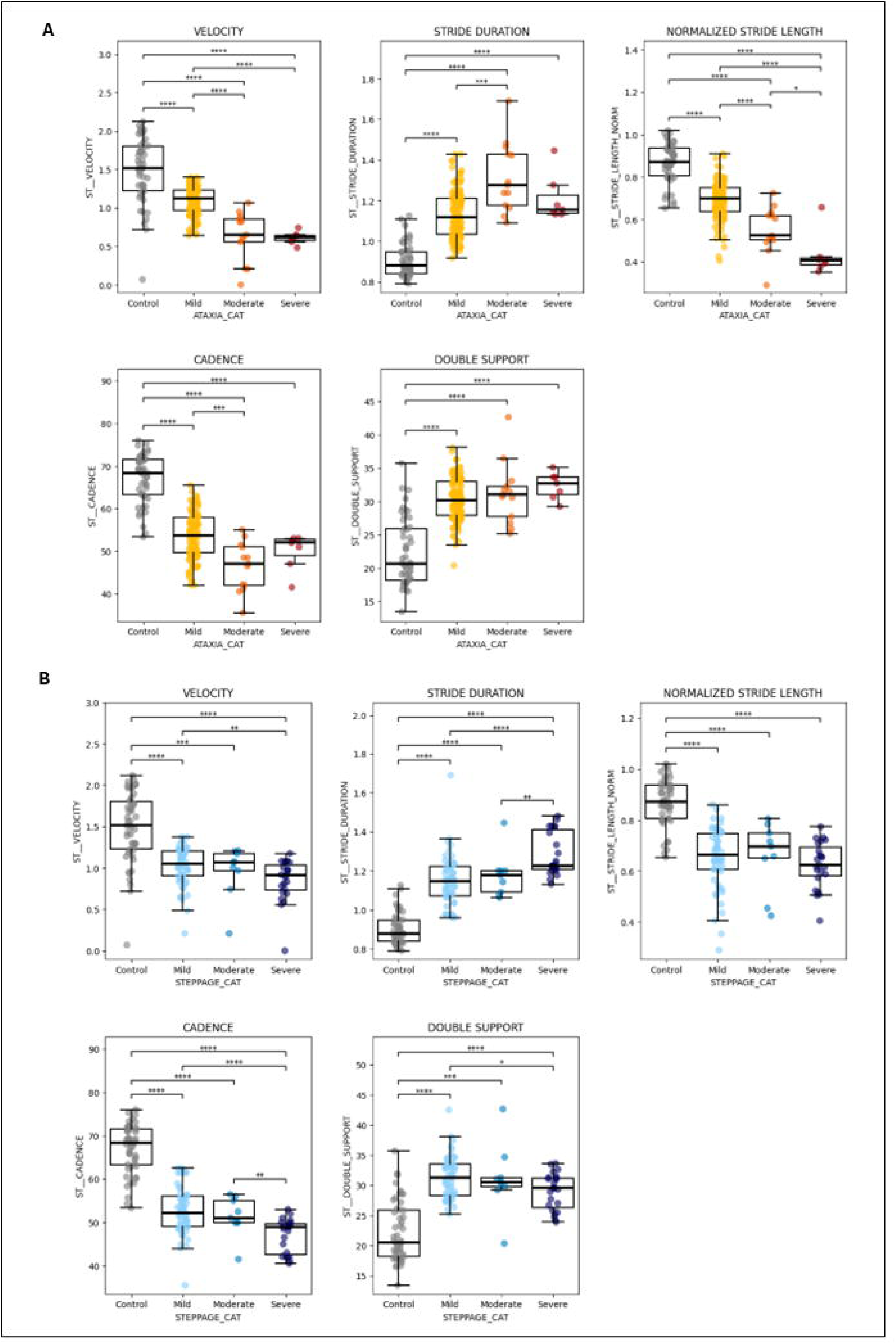
Analysis of spatiotemporalfeatures during one gait cycle. Velocity, stride duration, double support time, stride length, cadence and stride velocity represented for ataxia group (**A**) and steppage group (**B**). Kruskal-Wallis test performed, followed by post-hoc comparisons using Tukey’s HSD test. The differences between patients and controls have been represented as follows: **** ->p<0.0001. *** -> p<0.001. ** ->ρ<0.01. * -> ρ<0.05.

Plantar pressure or vertical force (VF) was analyzed throughout a gait cycle (Fig. 3A, 3B). For visual and analytical purposes, 3 features of the cycle were highlighted: the slope of the curve, the value in the valley, and the value of the second peak. In the group of patients with ataxia (Fig. 3A), the three gait cycle curves presented flatter slopes and significant differences than those of the control group. An inversion of the curve was also observed, with the second peak being higher (except for the severe ataxia group). For the steppage group (Fig. 3B), differences in slopes were not as prominent as those in the ataxia group, but an inversion of the second peak and an anticipation of this peak compared with those of the controls were also observed.

**Figure 3.**
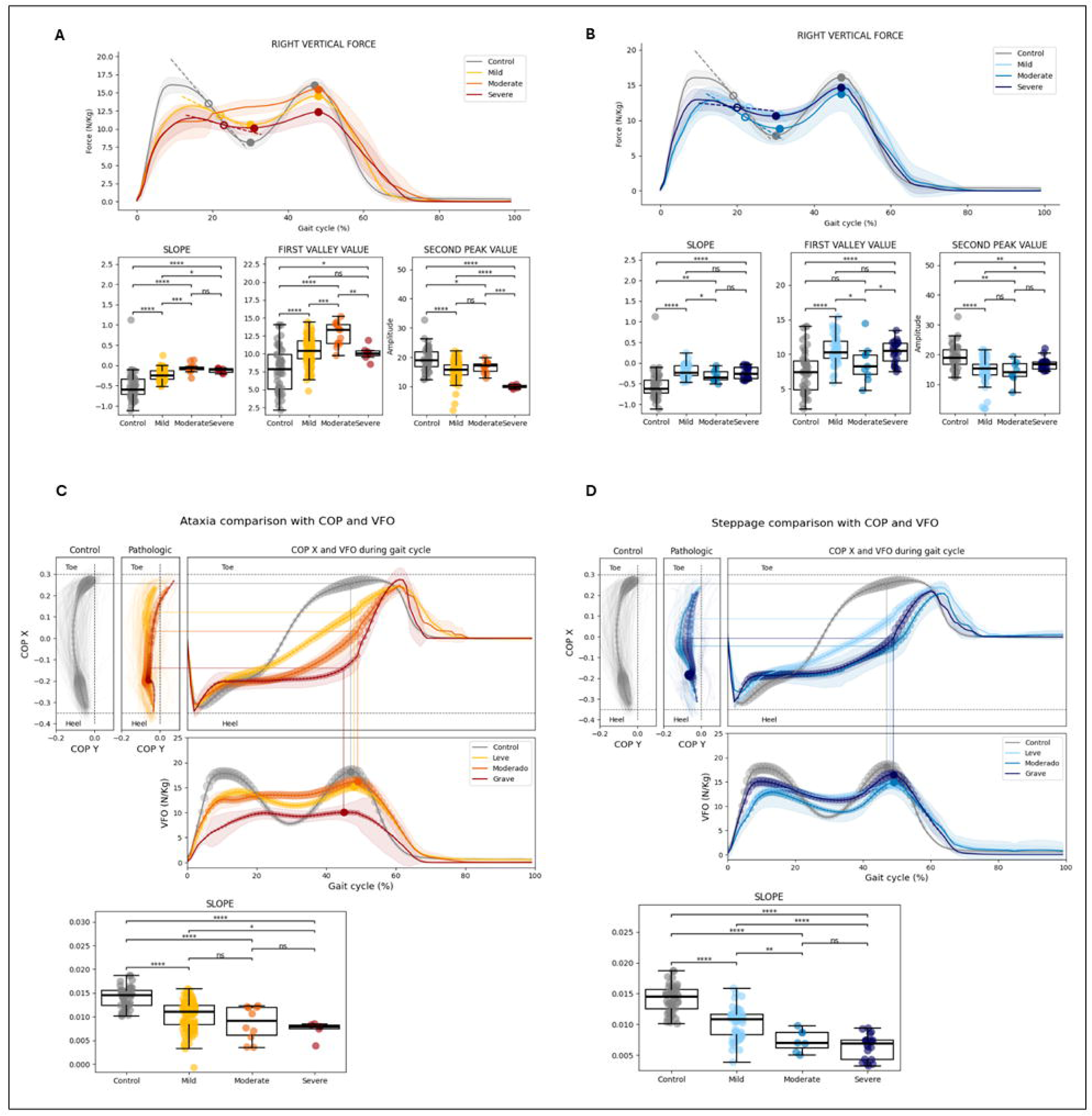
Results of vertical force (\T) and caitre of pressure (COP) during one gait cycle. Represented VFO for ataxia group (A) and for steppage group (B). Highlighted, slope, value of valley and value of the second peak features. Represented CO? for ataxia (Q and for steppage group (D). The size of the bubbles appearing on the curves represent the intensity of the vertical force during each moment of the gait cycle For both groups of patients highlighted the slope feature Kruskal-Wallis test performed. followed by post-hoc comparisons using Tukeys HSD test. The differences between patients and controls have been represented as follows: **** -> p<0.0001. *** -> p<0.001. ** -> p<0.01. * p<0.05.

The center of pressure (COP) of the vertical force over the insole during a gait cycle is displayed in Fig. 3C and 3D. In normal controls, the weight is distributed mainly between the heel, at the beginning of the cycle, and the toe, at the end of the cycle. In patients with ataxia (Fig. 3C), the weight is distributed on the heel and along the foot during the gait cycle, and there is almost no support of the weight with the toe. Additionally, a significant reduction in slope is detected across the different severity groups (except for the moderate and severe groups). Similar findings were observed for steppage patients (Fig. 3D), although differences across the severity groups were clearer. In patients with severe steppage, support almost exclusively happens on the heel. In addition, the value of the slope decreased significantly in the different severity groups except for the severe group compared with the moderate group.

We then analyzed foot angle flexion (Fig. 4A, 4B) and chose the value in the valley and the value at the second peak features for statistical comparisons. Patients with ataxia (Fig. 4A) presented a significant decrease in negative foot angle flexion across the different severity groups compared with the control group during the double support phase, represented by the valley value. The same occurred during the final phase of the cycle, which corresponded to the value in the second peak, in which we observed a significant decrease in positive foot angle flexion in the different severity groups compared with the control group. Compared with that of the controls, the angle of flexion of the negative foot angle decreased during the double support phase, and the angle of flexion of the positive foot angle also decreased during the final acceleration phase in the group of steppage patients (Fig. 4B). However, the differences between the severity groups were smaller than those between the ataxia groups.

**Figure 4.**
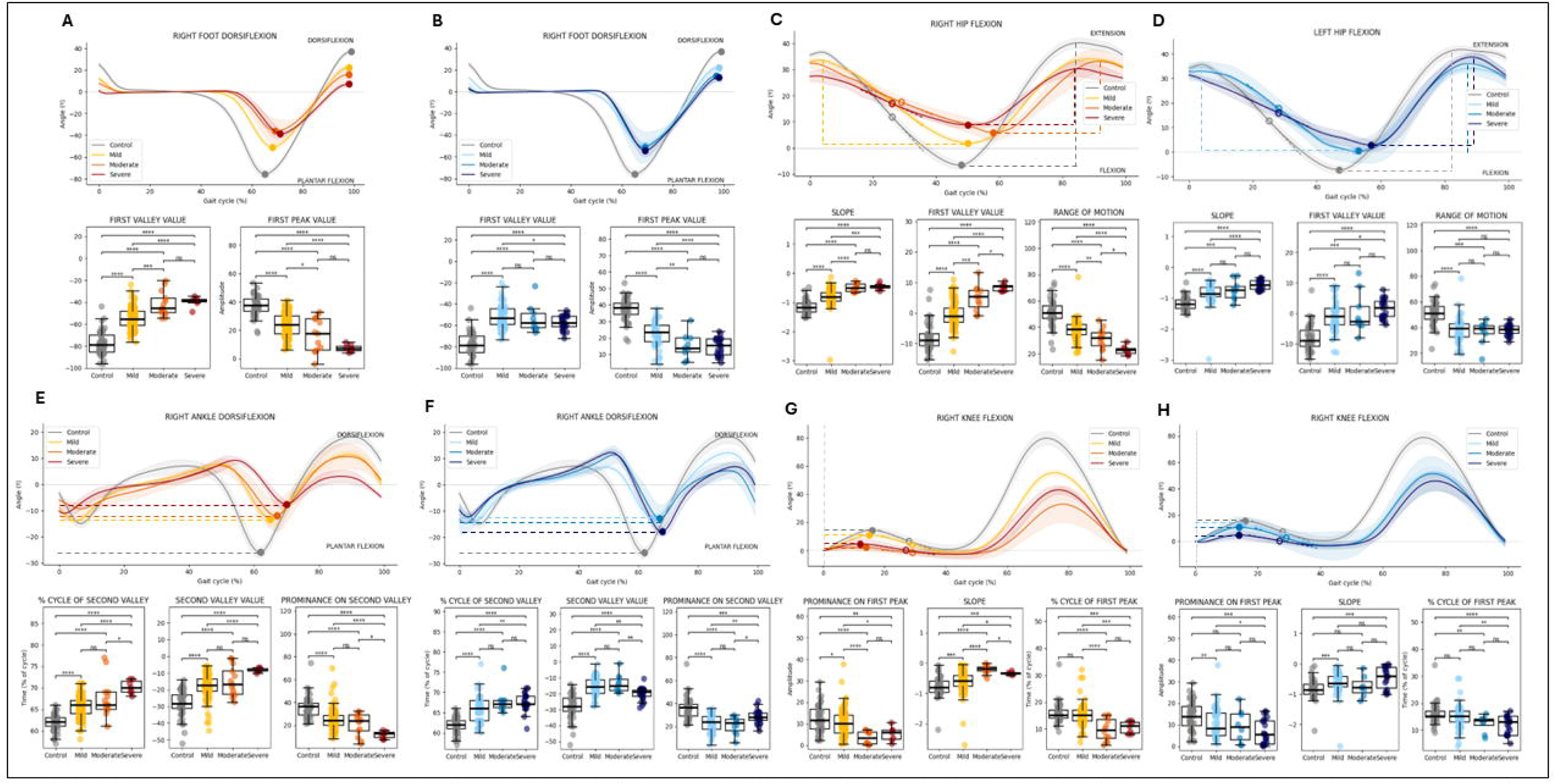
Results of foot angle flecion, hip angle flecion, ankle angle flecion, and knee angle flecion variables during one gait cycle. (**A, B**) For foot angle flexion variable, it was highlighted, valve of the valley and valve of the peak (**C D**) For hip angle flexion. it was highlighted slope, valve of the valley and range motion features. (**E, F**) Fer ankle angle flexion variable, it was highlighted, % cycle of second valley, valve of the second valley and prominence on second valleys in discontinued line. (**G, H**) For knee angle flextion, it was highlighted prominence on the first peak, slope and % cycle of first peakin discontinued line. Graphics A, C, E and F are from ataxia group and graphics B, D, F, H are from steppage group. Kruskal-Wallis test performed, followed by post-hoc comparisons using Tukey’s HSD test. The differences between patients and controls have been represented as followws: **** -> p<0.000l, *** -> p<0.001. ** -> p<0.01. * -> p<0.05.

The hip angle flexion variable (Fig. 4C, 4D) was associated with a decrease in slope and a significantly lower negative hip angle across the different severity groups of ataxia patients (Fig. 4C). The range of motion feature was also significantly lower in the different severity groups. For the steppage group (Fig. 4D), significant differences were observed in the value of the slope for the three severity groups compared with the control group. The values at the valley and the range of motion only significantly changed when patients and controls were compared but not across the three severity groups.

The ankle flexion variable (Fig. 4E, 4F) showed significant differences in the curves, indicating a delay in the appearance of the valley in patients with ataxia (Fig. 4E). A decrease was also observed in the negative value of the valley feature across the different severity groups compared with the control group. For this variable, the proportion of the cycle in the second valley significantly decreased across the different severity groups. The opposite occurred for the prominence feature, which increased significantly across the severity groups. For steppage patients (Fig. 4F), the same findings were observed: a delay in the appearance of the valley, a decrease in the negative value of the valley and in the % cycle of the second valley, and an increase in the prominence feature. The differences across severity groups were less prominent than those in the ataxia group.

For the knee angle flexion variable (Fig. 4G, 4H), significant differences were found for the ataxia group (Fig. 4G). A flattening of the entire first phase of the gait cycle was observed in patients, with a significant decrease in the slope (close to 0 across the different severity groups). The prominence and proportion of first peak features significantly decreased (except for those in the severe group) across the different severity groups. For the steppage group of patients (Fig. 4H), the same trends were observed: a decrease in the negative slope and a decrease in the prominence and proportion of first peak features. For the steppage group, significant differences were only obtained when comparing patients and controls and not across severity groups.

### DBB correlates with gait patterns in the global population of patients with PN

After the ability of the DBB system to capture deviations from normality in the different features was studied, the relationships of these features with their gait patterns were analyzed. The same cohort of patients was selected, and the LME test was used to analyze which biomechanical features correlated with each gait phenotype.

LME analysis revealed 219 biomechanical features associated with ataxia, steppage or both gait patterns (Supplementary Fig. 2). Of these, 131 features were associated with ataxia, 94 with steppage, and 133 with both gait patterns. Among these features, 24 were associated only with ataxia, and 10 were associated only with steppage. Interestingly, while the features associated with only gait ataxia were related mainly to plantar pressure and hip features, those correlated with steppage were related mainly to COP features. As previously observed in Fig. 3C and 3D, significant differences in COP features were observed among steppage patients and across the severity groups. Biomechanical features related to the foot, knee, ankle, and spatiotemporal variables correlated with both gait ataxia and steppage gait, in line with previous analyses shown in Fig. 2 and 4.

### DBB correlates with conventional objective and subjective clinical scales in the global population of patients with PN

For this second endpoint, the full cohort of patients was included, and the LME test was used to analyze which biomechanical features correlated with the different clinical scales (Supplementary Fig.3).

A total of 219 features were extracted, and 182 of them correlated significantly with any of the objective (MRCss, MICARS-SARA, INCAT and vigorimeter) or subjective (iRODS) clinical scales and the 2MWT. Four ST features, eleven-foot features, ten hip features, and seventeen knee features showed strong correlations with all the clinical scales and the 2MWT. Thirty-seven (17%) features did not correlate with any clinical scale. Other features related to VF, COP, hip and ankle correlated with some but not all the scales and 19 (8.7%) of them correlated with only one clinical scale or 2MWT. Focusing on the scales, iRODS correlated with 112 features (51.1%), MRCss with 100 features (45.7%), INCAT with 115 features (52.5%), MICARS with 96 features (43.8%), vigorimeter with 95 features (43.4%) and 2MWT with 147 features (67.1%) related to some of the VF, COP, knee, hip, and ankle features.

### DBB captures longitudinal changes in patients with neuropathies

We then studied the ability of the DBB to capture longitudinal changes in patients with PN. Patients selected for these analyses included those who presented with clinically meaningful worsening or improvement, defined as (1) a 2-point or greater change in the MRC score and/or (2) a 4-point change on the iRODS scale, which are conventional definitions for minimal clinically important differences in PN. We detected 41 CIDP patients (119 tests), 20 IgM-MGUS-associated neuropathies (54 tests), and 11 hereditary neuropathies (16 tests) that fulfilled these criteria and compared their results with those of a longitudinal assessment of 50 healthy controls.

When the best and worst clinical assessments in patients who showed a significant change in the MRCss scale were selected, significant differences were observed in 16 biomechanical features (Fig. 5), mainly those related to foot biomechanics. Additionally, differences were observed for COP variables and stride length features. Using the same statistical test but with a cohort of patients who presented a change of at least 4 points on the iRODS scale, we did not detect significant changes in any biomechanical feature, probably because the iRODS, a subjective scale, may fluctuate in the absence of objective change.

**Figure 5.**
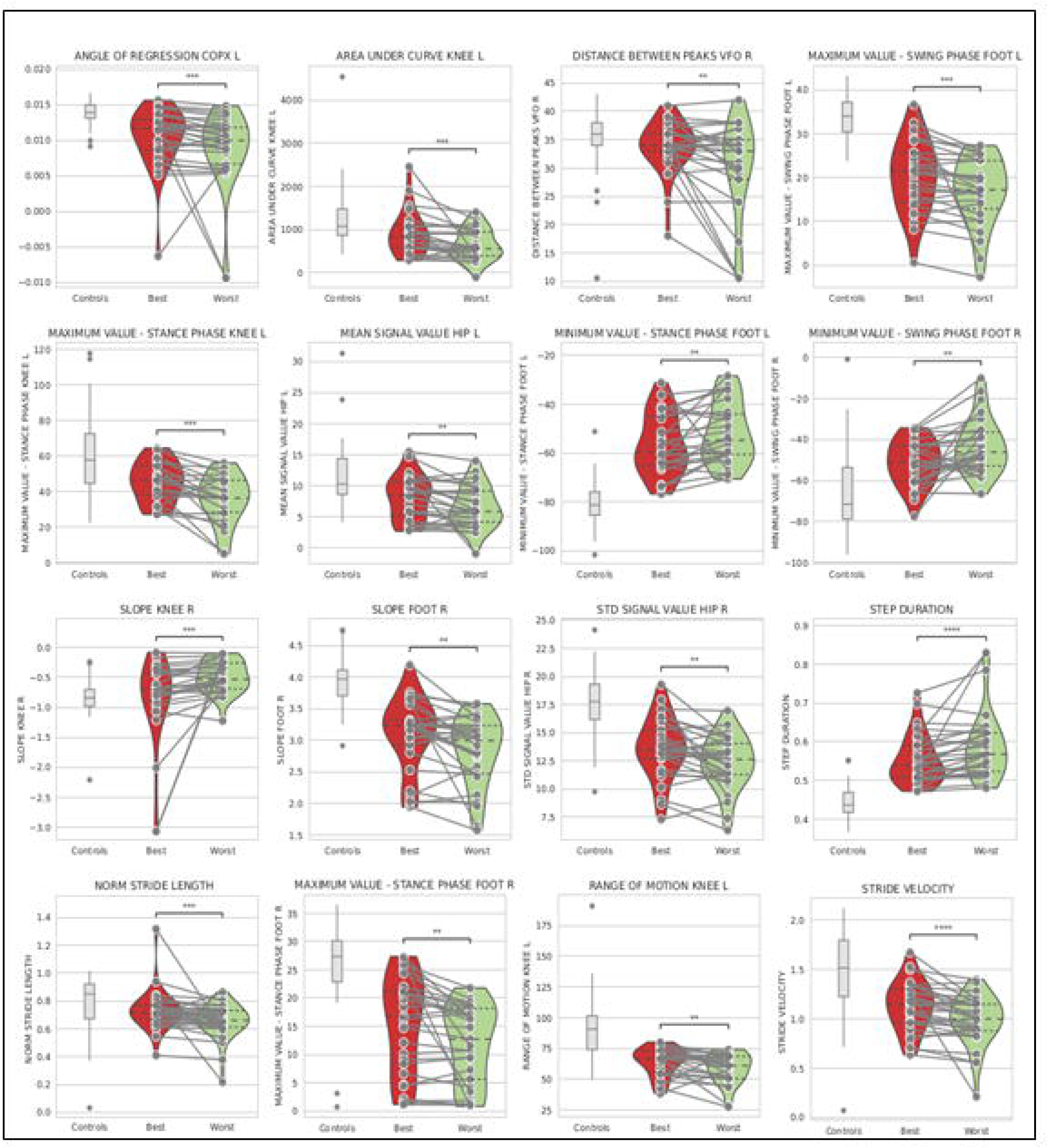
Fer>tureε that detected clinically significant changes in longitudinal analysis using Wilcoxon model. Best and the worst clinical assessments were selected in a cohort of patients who shewed a difference of. at least. 2 points on the MRCss. Multiple comparisons were used and a p value < 0.0C01 was stablished to differentiate the features which resulted significantly different over the tine. MRCss: Medical Research Council sum score.

When all available longitudinal assessments with LME analysis were used in a cohort of patients who presented a clinically relevant change in the MRCss at any time point, the DBB was able to identify 28 different biomechanical features correlated with clinical changes (Fig. 6), which were related to the foot, COP, ankle, and velocity. Finally, in a cohort of patients who had a change of 4 points in the iRODS, 37 biomechanical features correlated with the clinical change according to the same LME statistical test (Fig. 7). The features capturing the clinically relevant changes were those related to the foot. Significant longitudinal differences were also detected in features related to the hip and diverse spatiotemporal features in this cohort of patients. Ten of these features, four of which were related to foot function, detected the clinical changes classified by both the iRODS and MRCss (Fig. 6 and 7, marked in red*). Additionally, three of these features were related to knee function, two were related to spatiotemporal variables and one was related to hip function.

**Figure 6.**
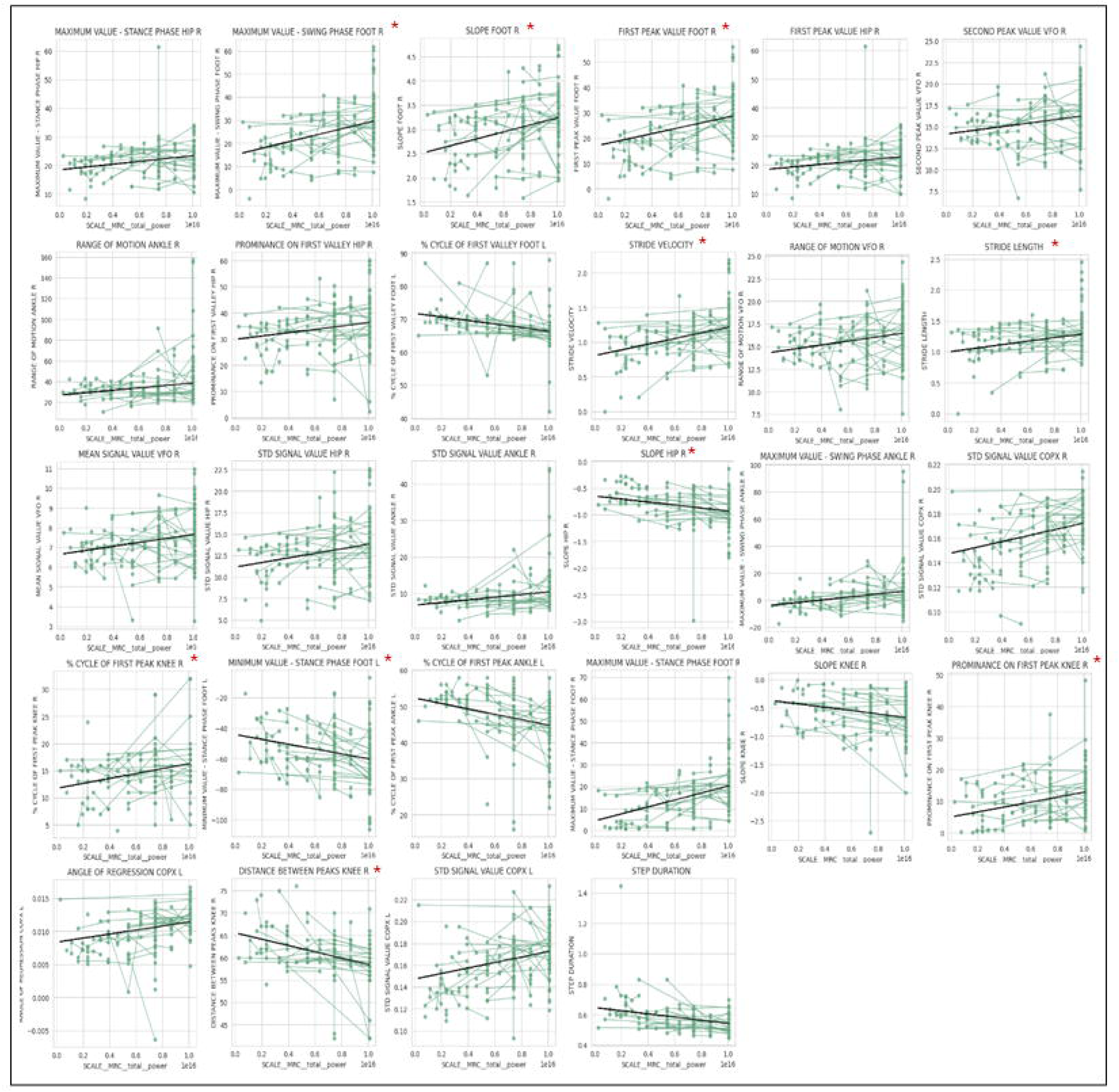
Features that detected clinically significant changes in longitudinal analysis using LME model. Used all available longitudinal assessments in a cohort of patients v.ho showed a clinical change of 2 points on MRCss scale. Multiple comparisons were used and a p value < 0.0001 was stablished to differentiate the features which resulted significantly different over the time. *Features that were also present in the cohort of patients who presented a clinical worsening in the iRODS scale. LME: linear mixed effects, MRCss: Medical Research Council sum score. iRODS: Inflammatory Rasch-built Overall Disability-Scale.

**Figure 7.**
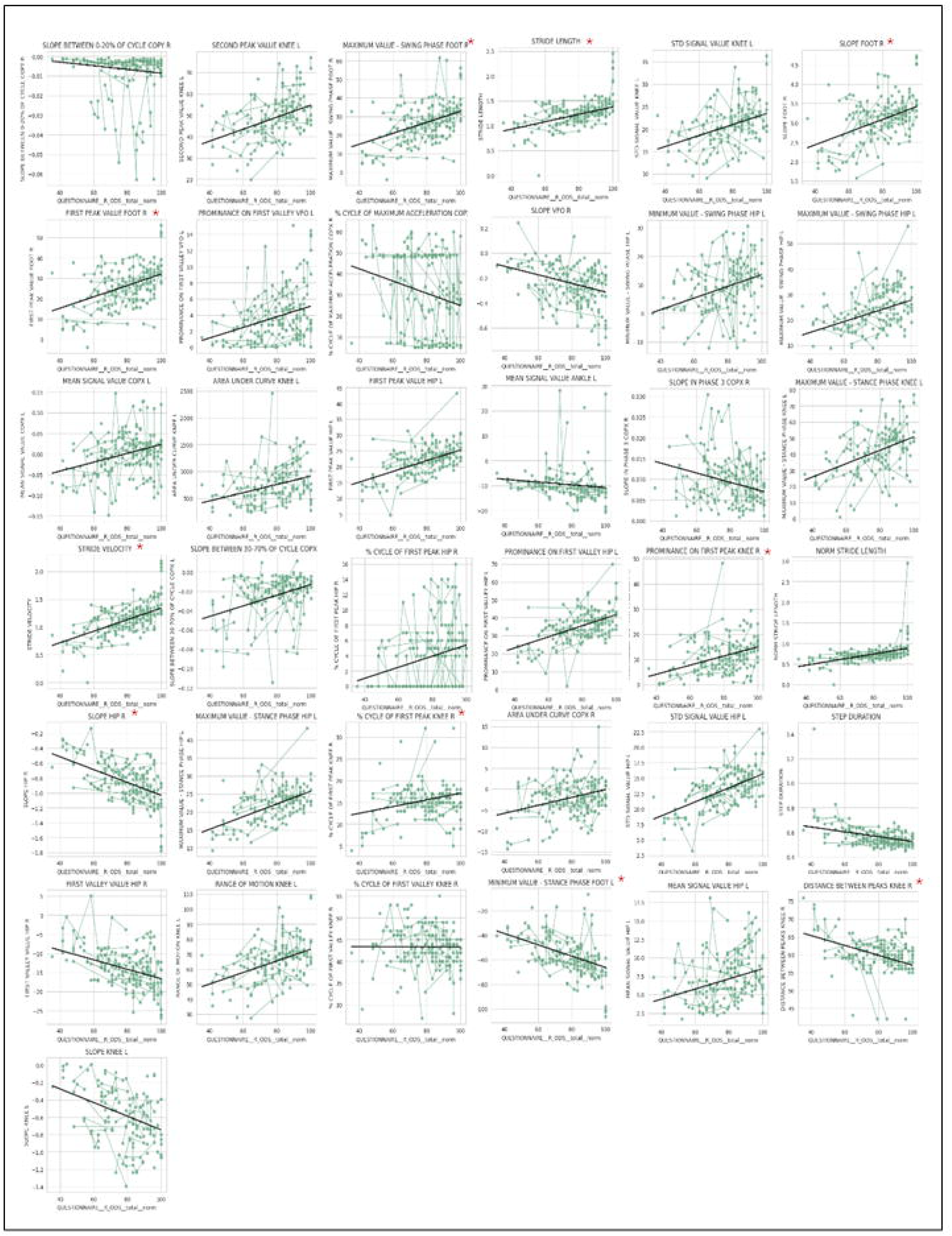
Fentures that detected clinically significant changes in longitudinal analysis using LME model. Used all available longitudinal assessments in a cohort of patients who shelved a clinical change of 4 points or grater in iRODS scale. Multiple comparisons were used and a p value < 0.0001 was stablished to differentiate the features which resulted significantly different over the time. *Features that were also present in the cohort of patients who presented a clinical worsening in the MRCss scale LME linear mixed effects. MRCss: Medical Research Council sum score. iRODS: Inflammatory Rasch-built Overall Disability-Scale.

## Discussion

Our study provides proof of concept that DBB objectively captures gait impairment and is responsive to clinical changes in patients with PN. DBB alterations correlate with PN patient disability, as measured with clinical scores. Moreover, longitudinal analysis of specific DBB features revealed clinical changes in patients with PN. This suggests that, if validated in appropriate clinical trials, DBB could be used to objectively monitor disease status and capture and quantify longitudinal changes in that clinical status in patients with PN. These objective and quantifiable assessments are achieved by integrating data signals captured with easy-to-use multiparametric sensors that can be simplified or tailored to select the most informative features in each type of gait phenotype.

A critical unmet need in the field of peripheral neuropathies is the absence of objective diagnostic, prognostic, and disease activity biomarkers^13^. Current monitoring strategies are based on clinical examinations and scores that are affected by subjective interpretation, imprecision, nonrelevant fluctuations and ceiling and floor effects. Ultimately, this results in substantial uncertainty regarding the clinical status of patients, particularly those with predominantly subjective symptoms, or those in which residual disability, treatment fluctuations and treatment side effects interfere with the proper evaluation of their clinical status^40^. Moreover, the subtle nature of clinical changes in hereditary neuropathies and the important placebo effect and the subjective and objective clinical fluctuations in patients with inflammatory neuropathies also influence the ability of clinical trials to detect significant changes, since the magnitude of the effect needs to overcome the substantial statistical noise that these imprecise scales provide. For these reasons, our main objective was to evaluate whether gait alterations (one of the main clinical manifestations in patients with PN) could be objectively assessed via a novel, multiparametric DBB that is able to capture and quantify the biomechanical alterations associated with gait impairment.

We first proved that DBB is able to detect differences in two different gait phenotypes commonly observed in patients with PN (ataxia and steppage gait) and across severity groups. Second, we demonstrated that some DBB alterations correlate with conventional clinical scales and PROMS. Finally, we validated the ability of DBB to detect changes in the different features over time.

Gait impairment is one of the most characteristic alterations in patients with PN. As such, one of the most obvious alterations in the clinical assessment of gait is a decrease in gait speed. For this reason, timed walking tests are popular in the assessment of PN and other neuromuscular disorders. However, the exact parameters that determine a reduction in gait speed differ across patients, and there may be subtle alterations in specific parameters even if, globally, gait is not visually impaired or if the distance achieved during the timed test is considered within normal ranges. Considering the frequent alteration of gait speed in patients with PN, we consequently observed that spatiotemporal parameters related to gait speed, such as velocity, stride length and cadence, decreased significantly in these patients; in addition, the parameters related to time increased significantly with severity. Although these alterations in spatiotemporal gait features are detected in both ataxia and steppage phenotypes and strongly correlate with all clinical scores, the specific features that are altered and the pattern of abnormality differ depending on the type of gait. Essentially, patients with ataxia or steppage walk slower than normal controls do, but the features behind this decrease in speed are different in each gait phenotype, and the degree of alteration of each spatiotemporal feature varies depending on the severity of the disease.Similarly, gait impairment in patients with PN also results in footprint abnormalities that vary depending on gait phenotype. Consequently, alterations in plantar pressure or vertical force allowed us to distinguish both groups of patients whose gait cycle curves were significantly different from each other. This parameter clearly captured significant differences across the different levels of severity, especially in the ataxia group. In the severe ataxia group of patients, the inversion of the second peak was lost because these patients presented high gait instability, and both feet barely lifted off the ground. However, the double support phase was not impaired to the same extent in steppage patients. Footprint abnormalities are visually obvious in patients with gait alterations, but our study again provides objective and quantifiable evidence of these alterations and detects which features are specific for each type of gait.

In steppage patients, the main gait alteration is foot drop. This finding suggests that plantar pressure is likely significantly disrupted in patients whose foot control is significantly impaired. As such, COP features (in which the plantar distribution of pressure that the foot exerts when touching the ground is evaluated) were the only features that showed a significant correlation with the steppage gait pattern. COP features on the step page were significantly disrupted and differed among the three severity groups and compared with those of the control group. These results mirror what is observed in the clinic, where patients whose foot and toe drop do not exert plantar pressure in the anterior regions of the insole pressure detectors. However, although differences were observed between ataxia patients and controls, there were no differences across severity groups.

The foot, hip, ankle, and knee angle flexions also exhibited notable deviations from normal in the different patient groups. Patients with ataxia show a pattern of gait that results from the presence of imbalance, whereas steppage gait appears due to the presence of distal leg weakness. Since steppage gait abnormalities are more focally distributed, deviations from the normality of leg angle features of the DBB were more generalized and profound in patients with ataxia. For these reasons, foot flexion angle features were altered in both groups of patients, whereas hip angle flexion appeared flattened and correlated with severity in the ataxia group only. Patients with steppage do not show flat curves in hip flexion because, when their foot is dropped, they exert additional force from the hip to be able to lift the entire foot, preserving or even exaggerating the width of the hip angle. In contrast, ataxic patients tend to walk without bending their hips and knees or widening the sustentation base to increase stability and avoid falls. Ankle and knee flexion feature alterations were more profound, as were other angle flexion features, for ataxia patients. Leg joint angle features made it possible to stratify the different severity groups in ataxia patients but not in steppage patients, in which differences were only able to distinguish patients from controls.

Assessing gait with DBB would only be relevant if the findings correlate with validated clinical scales. In the group of diseased patients, the 2MWT and all objective and subjective clinical scales had strong correlations with DBB features related to the foot and with some of the temporospatial variables. The 2MWT, the test used for all data analysis, proved to be the test, as expected, that correlated with more biomechanical features. Some biomechanical features did not show correlations for any clinical scale or for the 2MWT. Only grip strength, which was assessed with a vigorimeter and the iRODS, a disability scale validated to capture the full range of disability in inflammatory neuropathies, also showed significant correlations with vertical force features; COP features; and ankle, hip and knee angles, such as the MRCss, MICARS-SARA and INCAT scales. Some of these correlations may be obvious and visually detected since increased severity of a neuropathy will certainly correlate with spatiotemporal features. However, this exhaustive analysis helps uncover other, less obvious correlations, such as those between grip strength and COP features. Overall, these results indicate that biomechanical alterations correlate with the clinical status of patients and, thus, could be a way to objectivize and quantify deviations from normality.

To be clinically useful for monitoring disease in clinical practice or in clinical trials, DBB needs to be able to capture longitudinal changes in addition to correlating with clinical scales and capturing different grades of severity. To initially test the ability of DBB to capture longitudinal changes, we selected patients for whom a clinically significant change was defined by 2 points on the MRCss scale or 4 points on the iRODS scale, which are two frequently used criteria to detect a minimal clinically important difference and analyze their DBB features. Several DBB features were able to detect longitudinal changes over time: when all available data were used, 28 and 37 features detected clinical changes defined with MRCss and iRODS, respectively. Most of them are related to foot features because, when weakness appears in patients with PN, it most frequently starts in the most distal muscles. However, if, instead of considering all available tests, we selected the best and worst clinical assessments in these two cohorts of patients, 16 DBB features became significant for the MRC group, suggesting that DBB monitoring is not only able to detect minimal clinical longitudinal changes but also that when the disease progresses further, as expected, other DBB features become sensitive to those changes. These results demonstrate that the DBB features most sensitive for detecting longitudinal changes are those related to the foot. This can certainly have implications for setting the minimum number of parameters that are clinically useful so that gait analysis can be simplified. There were no significant changes in any biomechanical features when the clinical status was stratified with the iRODS. This is likely because the changes detected by the iRODS scale are more subtle and influenced by the patient’s subjectivity.

There are several limitations for multiparametric wearable technology that need to be acknowledged. From a feasibility point of view, the duration of the visits that incorporate DBB analysis is significantly longer than that of conventional visits since all sensors need to be placed and synchronized. This limitation could be mitigated by choosing a set of sensors that provides sufficient information to capture clinically relevant alterations across groups and longitudinally within groups. Another option is to restrict the use of the technology to research settings, for example, in clinical trials, in which additional time can be allocated to use these technologies. Additionally, handling and interpreting massive amounts of biomechanical data is challenging. In the future, the simplification of the protocol, linked to the development of clinically validated software that simplifies the interpretation of the findings, would be necessary to be able to incorporate the technology into the clinical routine.

Our study also has several limitations. First, and most importantly, a diverse set of patients with neuropathy, with different clinical features, different severities, and different longitudinal behaviors, is incorporated. Although we believe that this approach is interesting if we want to find biomechanical features that are useful for monitoring any PN patient, regardless of type, focused studies stratifying each disease type could provide more precise DBB features for use in each neuropathy subtype. Nevertheless, the fact that we were able to capture DBB features correlated with clinical status and longitudinal changes despite the heterogeneity of the population suggests that a disease-focused approach would further increase the sensitivity of this technology. Future analyses should consider stratification of findings according to disease instead of stratification according to gait phenotype. Another limitation of the study is that DBB is only useful when gait impairment is present. Although this is a very frequent feature in patients with neuropathy, it is not universal. Future studies should address whether this technology is also able to capture minor DBB alterations in patients whose gait disturbances are not obvious. Another limitation of the study is that these results are applicable only to adult populations. How DBB alterations behave in children with neuropathies and if those are comparable to findings in adults is a pending task. Finally, all these results arise from the interpretation of individual DBB feature data. An important step is to develop a paradigm of analysis in which the interplay of diverse DBB features and clinical scores can provide a more precise picture of the clinical situation of a patient.

## Conclusion

Our study provides proof-of-concept that noninvasive wearable biomechanical sensors are useful for capturing, quantifying and monitoring gait disturbances in patients with PN in an objective and reproducible way. Future multicenter clinical trials should address if DBB is reproducible, if stratification according to disease type improves its performance and if it is able to detect subtle alterations in patients in which gait disturbances are not visually obvious. Successful validation of the DBB system will help overcome the limitations of current monitoring strategies by providing more objective and precise monitoring in PN that helps optimize patient care and, ultimately, improve the quality of life of these patients.

## Data availability

The datasets generated and/or analyzed during the current study are available from the corresponding author upon reasonable request.

## Acknowledgements

The authors acknowledge the Department of Medicine at the Universitat Autonòma de Barcelona. The authors also thank all our patients for their support and collaboration. C.T.I, M.C.A, L.M.A, E.P.G, R.C.V, A.V and A.C are members of the European Reference Network for Neuromuscular Diseases – Project ID N° 870177.

