## Supplementary material for "Digital biomechanical biomarkers in the clinical assessment of patients with peripheral neuropathies"

| | Ataxia<br>MICARS-SARA item1<br>(walking capacities) | Steppage<br>MRC-SS Ankle Dorsiflexion<br>item (Right + Left) |
|---|---|---|
| **Severe** | Value ≥ 5 | Value ≤ 4 |
| **Moderate** | Value 3-4 | Value 5-6 |
| **Mild** | Value 1-2 | Value 7-8 |
| **Normal** | Value 0 | Value ≥ 9 |

**Supplementary Table 1. Gait pattern classification in ataxia or steppage and according to de severity.** Categorization performed following the score obtained in item 1 of MICARS-SARA scale for ataxia patients and following the score of dorsiflexion item for steppage patients. MICARS-SARA: Modified Internacional Cooperative Ataxia Rating Scale and Scale for the Assessment and Rating of Ataxia; MRCss: Medical Research Council sum score.

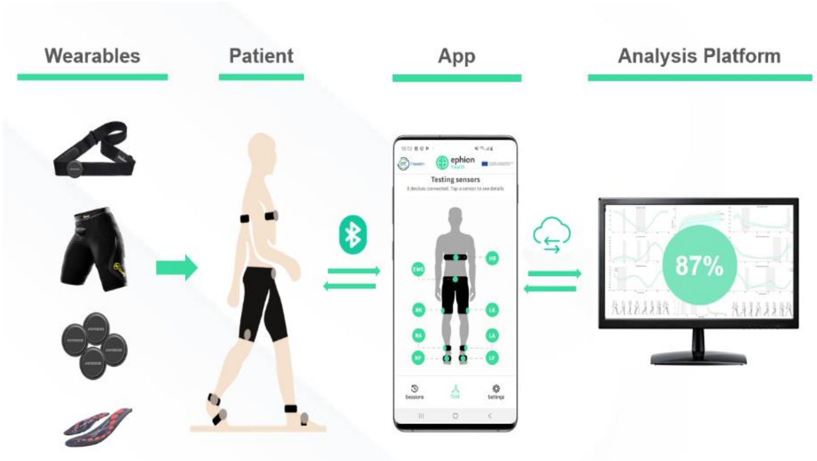


**Supplementary Figure 1. Ephion Mobility System.** It is made up of 5 inertial sensors located in different parts of the body, pants with surface EMG and insoles with inertial and plantar pressure sensors. Using Bluetooth these sensors are synchronized into smartphone and to a platform to perform the data analysis.

| | Gait pattern | | N |
|---|---|---|---|
| | **Steppage** | **Ataxia** | |
| | Mild | Mild | 17 |
| | | Moderate | 2 |
| | | Severe | - |
| | | Normal | 4 |
| | Moderate | Mild | 3 |
| | | Moderate | 1 |
| | | Severe | 1 |
| | | Normal | - |
| | Severe | Mild | 4 |
| | | Moderate | 2 |
| | | Severe | - |
| | | Normal | - |
| | Normal | Mild | 16 |
| | | Moderate | 1 |
| | | Severe | 1 |
| | | Normal | 26 |
| **Not specified** | - | - | 1 |

**Supplementary table 2. Patients included in the study according to their gait pattern.** Number of patients with ataxia and/or steppage and patients with normal gait pattern. Patients with normal gait pattern were excluded for the analysis of normal and abnormal gait pattern whereas in the study of correlations with gait patterns these patients were included.

| | | Missing | Grouped by ATAXIA | | | | | |
|---|---|---|---|---|---|---|---|---|
| | | | Mild | Moderate | Severe | Normal gait | Controls | p-value |
| **N** | | | 40 | 6 | 2 | 30 | 50 | |
| **Age (years), mean (SD)** | | 0 | 62.3 (12.7) | 67.4 (10.4) | 57.4 (5.8) | 58.7 (13.5) | 59.9 (9.4) | 0.360 |
| **Sex, male (%)** | | | 29 (72.5) | 3 (50.0) | 1 (50.0) | 18 (60.0) | 17 (34.0) | |
| **2 MWT distance (m), mean (SD)** | | 0 | 124.8 (26.2) | 54.0 (40.4) | 79.0 (15.6) | 158.2 (28.7) | 198.0 (35.3) | <0.001 |
| **Vigorimeter, mean (SD)** | Left | 50 | 67.8 (22.4) | 45.5 (13.3) | 26.0 (19.8) | 77.8 (27.0) | NA | <0.002 |
| | Right | 50 | 66.8 (21.1) | 45.7 (11.1) | 54.0 (22.6) | 71.9 (27.3) | NA | 0.081 |
| **MRC total, mean (SD)** | | 50 | 57.6 (2.4) | 51.8 (4.2) | 53.5 (3.5) | 59.1 (1.4) | NA | <0.001 |
| **INCAT total, mean (SD)** | | 51 | 2.0 (1.1) | 4.2 (1.2) | 5.0 (0.0) | 1.6 (1.2) | NA | <0.001 |
| **iRODS total, mean (SD)** | | 58 | 36.3 (6.4) | 25.2 (6.5) | 21.5 (2.1) | 41.5 (5.3) | NA | <0.001 |

**Supplementary table 3. Baseline clinical characteristics according to the severity in ataxia group.** Number, mean, standard deviation (sd) of patients included of each pathology according to age and sex. Mean and sd results of 2-minute-walking-test (2MWT), grip strength using vigorimeter, Medical Research Council sum score (MRCSS), Inflammatory Neuropathy Cause and Treatment (INCAT), Inflammatory Rasch-built Overall Disability Scale (iRODS). Chi-quadrat test used for study the differences between sex. ANOVA test used to study the differences between each severity group for the clinical scales and 2MWT. A significance level of p-value <0.0001 established to differentiate these patient groups.

| | | Missing | Grouped by STEPPAGE | | | | | |
|---|---|---|---|---|---|---|---|---|
| | | | Mild | Moderate | Severe | Normal gait | Controls | p-value |
| **N** | | | 23 | 5 | 6 | 44 | 50 | |
| **Age (years), mean (SD)** | | 0 | 58.4 (14.8) | 60.5 (13.6) | 58.9 (15.1) | 63.1 (11.3) | 59.9 (9.4) | 0.954 |
| **Sex, male (%)** | | | 14 (60.9) | 3 (60.0) | 3 (50.0) | 31 (70.5) | 17 (34.0) | |
| **2 MWT distance (m), mean (SD)** | | 0 | 116.8 (33.0) | 99.0 (41.1) | 85.8 (50.0) | 148.2 (32.5) | 198.0 (35.3) | <0.001 |
| **Vigorimeter, mean (SD)** | Left | 50 | 64.3 (22.0) | 55.8 (30.9) | 55.0 (27.1) | 74.6 (26.1) | NA | 0.114 |
| | Right | 50 | 62.5 (20.6) | 60.4 (15.3) | 57.0 (28.4) | 71.2 (25.3) | NA | 0.312 |
| **MRC total, mean (SD)** | | 50 | 56.9 (2.0) | 53.8 (3.0) | 51.5 (3.5) | 59.3 (1.3) | NA | <0.001 |
| **INCAT total, mean (SD)** | | 51 | 2.4 (1.5) | 3.5 (1.3) | 2.7 (1.6) | 1.7 (1.2) | NA | 0.023 |
| **iRODS total, mean (SD)** | | 58 | 34.8 (8.0) | 29.8 (11.3) | 35.5 (7.2) | 39.2 (6.6) | NA | 0.028 |

**Supplementary table 4. Baseline clinical characteristics according to the severity in steppage group.** Number, mean, standard deviation (sd) of patients included of each pathology according to age and sex. Mean and sd results of 2-minute-walking-test (2MWT), grip strength using vigorimeter, Medical Research Council sum score (MRCSS), Inflammatory Neuropathy Cause and Treatment (INCAT), Inflammatory Rasch-built Overall Disability Scale (iRODS). Chi-quadrat test used for study the differences between sex. ANOVA test used to study the differences between each severity group for the clinical scales and 2MWT. A significance level of p-value <0.0001 established to differentiate these patient groups.

| Analysis of differences of normal and abnormal gait patterns | | | | | | | |
|---|---|---|---|---|---|---|---|
| **Gait pattern** | **Level** | **CANOMAD** | **CIDP** | **Hereditary neuropathy** | **IgM-MGUS** | **AN** | **N Total** |
| **Control** | **-** | - | - | - | - | - | 50 |
| **Normal** | **-** | 4 | 50 | 2 | 19 | 3 | 78 |
| **Ataxia** | **Mild** | 3 | 55 | 8 | 38 | 14 | 118 |
| | **Moderate** | 5 | 4 | 2 | 1 | 1 | 13 |
| | **Severe** | - | 7 | - | - | - | 7 |
| **Steppage** | **Mild** | 1 | 28 | 12 | 10 | 3 | 54 |
| | **Moderate** | - | 5 | 3 | - | 1 | 9 |
| | **Severe** | - | 11 | 1 | 3 | 8 | 23 |

| Longitudinal analysis | | | | | | |
|---|---|---|---|---|---|---|
| | **CANOMAD** | **CIDP** | **Hereditary neuropathy** | **IgM-MGUS** | **AN** | **N Total** |
| **Control** | - | - | - | - | - | 50 |
| **Change of ≥2 on MRCss** | - | 83 | 7 | 23 | 8 | 121 |
| **Change of ≥4 on RODS** | - | 56 | 2 | 29 | 6 | 93 |

**Supplementary Table 5. Number of tests included in the analysis of the differences between normal and abnormal gait pattern and in the longitudinal analysis.** Classification according to their diagnosis, gait pattern and severity in the case of the analysis of differences in gait patterns. Classification according to their diagnosis and to the change in total score in MRCss or iRODS scale. Disease classification mainly included the inflammatory neuropathies CIDP, IgM-MGUS, CANOMAD, another AN and hereditary neuropathies. CANOMAD: chronic ataxic neuropathy, ophthalmoplegia, immunoglobulin M [IgM] paraprotein, cold agglutinins, and disialosyl antibodies; CIDP: chronic inflammatory demyelinating polyneuropathy; IgM-MGUS: monoclonal gammopathy of undetermined significance associated with IgM; AN: autoimmune nodopathy; MRCss: Medical Research Council sum score; iRODS: Inflammatory Rasch-built Overall Disability Scale.

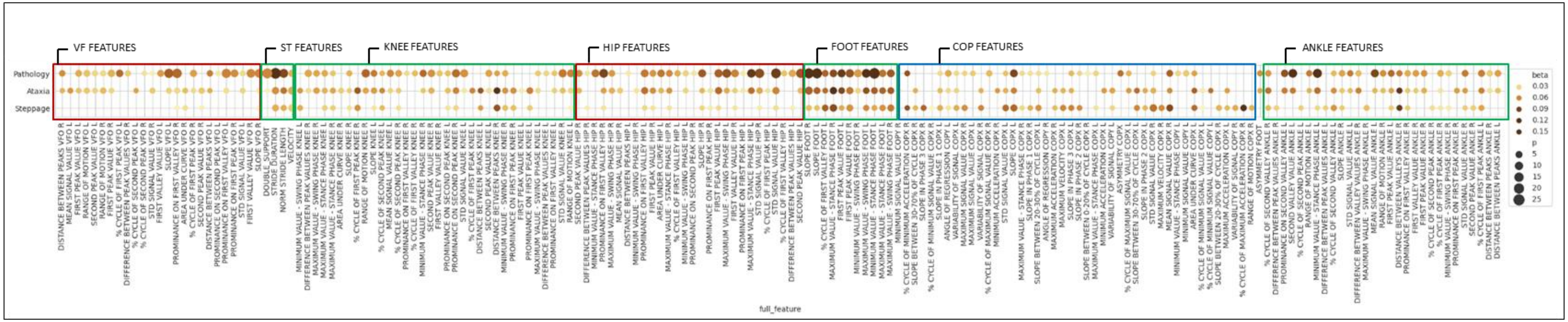


**Supplementary Figure 2. Correlation of gait patterns with biomechanical features.** Colour intensity and size of the bubbles represent the correlation level of the features with their gait pattern. Boxed in red, features related to ataxia gait pattern; boxed in blue, features related to steppage gait and boxed in green, features related to both gait patterns. Multiple comparisons were performed and a significance level of p-value <0.05 was established to select features that correlated with their gait pattern. Only plotted significant features. VF: Vertical force; ST: Spatiotemporal; COP: Center of pressure.

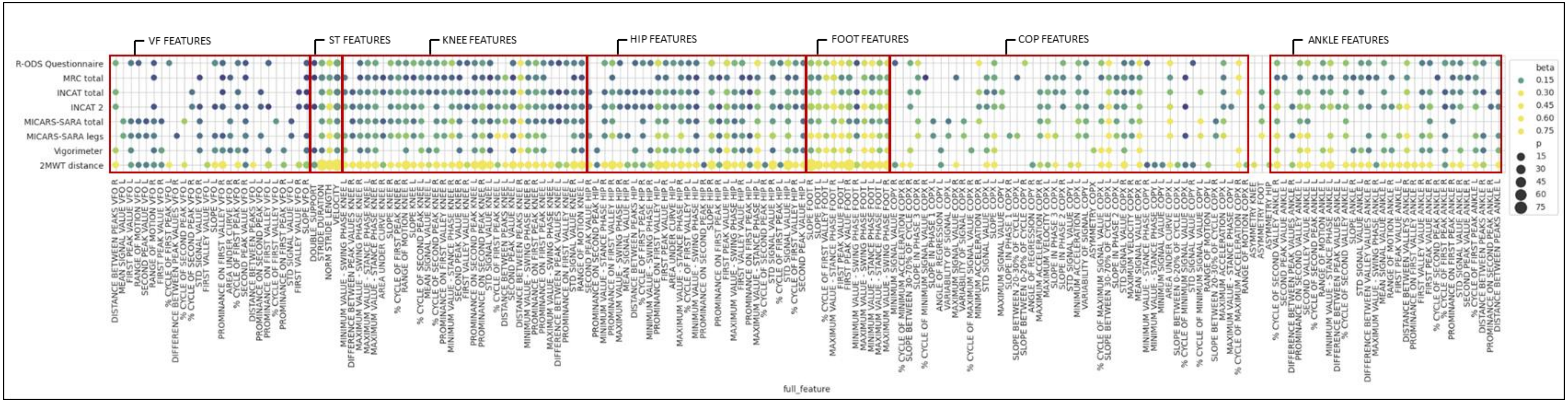


**Supplementary Figure 3. Correlation of clinical scales with biomechanical features.** Colour intensity and size of the bubbles represent the correlation level of the features with their gait pattern. Non-normalized and normalized scales are represented using the logarithm of the total score. Multiple comparisons were performed and a significance level of p-value <0.05 was established to select features that correlated with their gait pattern. Only plotted significant features. VF: Vertical force; ST: spatiotemporal; COP: Center of pressure; MRCss: Medical Research Council sum score, INCAT: Inflammatory Neuropathy Cause and Treatment, iRODS: Inflammatory Rasch-built Overall Disability Scale, MICARS-SARA: Modified Internacional Cooperative Ataxia Rating Scale and Scale for the Assessment and Rating of Ataxia, 2MWT: 2-minute walking test.

| Sensors | Variables | Features | Definition | Representation |
|---|---|---|---|---|
| **Inertial sensor (Movesense)** | Hip angle flexion 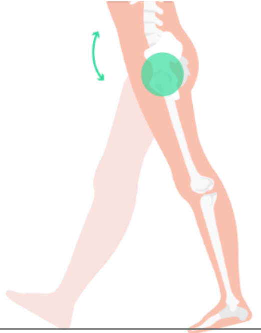 | ***Slope*** (1) | Slope value between maximum and minimum flexion-extension point during stance phase. | 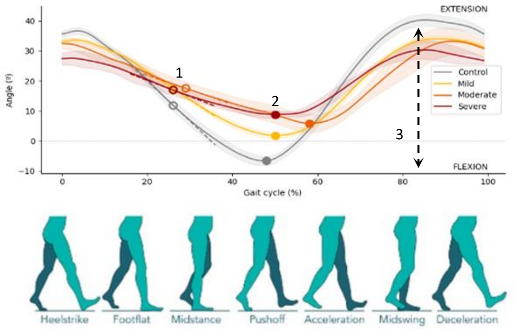  |
| | | ***First peak value*** (amplitude, ***time*** (%) and **prominence)** | Maximum point of flexion-extension at stance phase. Value and % of gait cycle when it occurs. | |
| | | **Second peak value** (amplitude, **time** (%) and **prominence**) | Maximum point of flexion-extension at swing phase. Value and % of gait cycle when it occurs. | |
| | | ***First valley value*** (2) (amplitude, **time** (%) and ***prominence***) | Minimum point of flexion-extension between middle-stance and middle-swing phase. Vale and % of gait cycle when it occurs. | |
| | | Range of motion (3) | Range of motion and distance in % of cycle between maximums and minimums flexion-extension points obtained. | |
| | Knee angle flexion 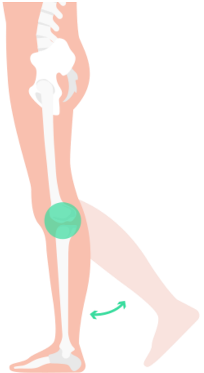 | ***Slope*** (1) | Slope value between maximum and minimum flexion-extension point during stance phase | 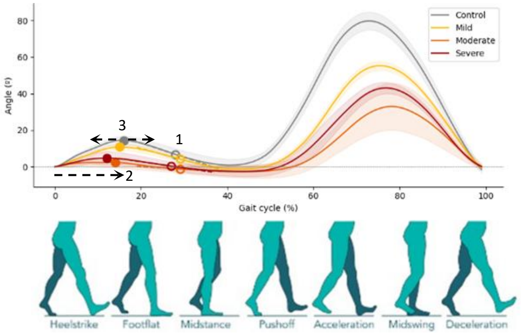  |
| | | **Maximum stance** (amplitude, **time** (%) and prominence) | Maximum point of flexion-extension between heel-off and middle-swing phase. Value and % of cycle when it occurs. | |
| | | First peak value (amplitude, ***time*** (%) (2) and ***prominence*** (3)) | Maximum point of flexion-extension between heel-strike and heel-off phase. Value and % of cycle when it occurs. | |
| | | ***Second peak value*** (amplitude, **time** (%) and **prominence**) | Maximum point of flexion-extension during swing phase. Value and % of gait cycle when it occurs. | |
| | | **First valley *value*** (amplitude, time and **prominence**) | Minimum point flexion-extension between middle-stance and toe-off phase. Value and % of gait cycle when it occurs. | |
| | Ankle angle flexion 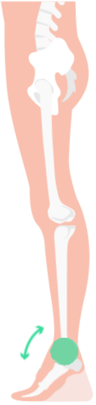 | **Slope** | Slope value between maximum and minimum flexion-extension point during stance phase | 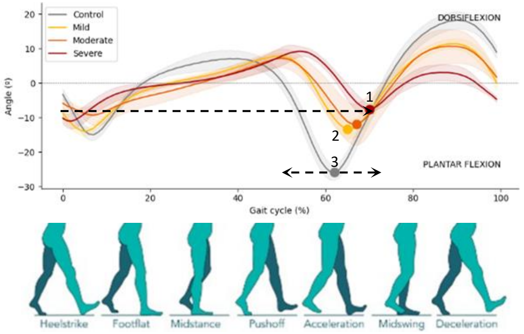  |
| | | **First peak value** (amplitude, ***time*** (%) and prominence) | Maximum point of flexion-extension between middle-stance and toe-off phase. Value and % of gait cycle when it occurs. | |
| | | **Second peak value** (amplitude, time (%) and **prominence**) | Maximum point of flexion-extension during swing phase. Value and % of gait cycle when it occurs | |
| | | **First valley value** (amplitude, time (%) and prominence) | Minimum point of flexion-extension during heel-strike. Value and % of gait cycle when it occurs. | |
| | | **Second valley value** (1) (amplitude, **time** (%) (2) and **prominence** (3)) | Minimum point of flexion-extension between pre-swing and middle-swing phase. Value and % of gait cycle when it occurs | |

| | | | | |
|---|---|---|---|---|
| | All inertial sensor signals common features | ***Flexion-extension distance between maximum and minimum flexion-extension values*** | Range of motion and distance in % of cycle between maximums and minimums flexion-extension points obtained. | |
| | | Flexion-extension slope during stance phase | Slope value between maximum and minimum flexion-extension point during stance phase | |
| | | ***Mean of flexion-extension signal*** | Mean value of flexion-extension during gait cycle. | |
| | | Flexion-extension variability | Standard deviation of flexion-extension during gait cycle. | |
| | | ***Flexion-extension area*** | Area under the curve of flexion-extension during gait cycle. | |
| | | Flexion-extension range of motion | Range of motion of flexion-extension during gait cycle. | |
| | | **Minimum and *maximum flexion-extension at* swing and *stance phase*** | Maximum and minimum point of flexion-extension during swing and stance phase. Value and % of gait cycle when it occurs. | |
| | | ***Distance* and amplitude *between peaks* and valleys** | Distance in % of cycle between maximums and minimums during gait cycle. Amplitude of the pattern between maximums and minimums during gait cycles. | |
| **Insoles (Moticon)** | Foot angle flexion 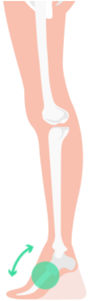 | ***Slope*** | Slope value between maximum and minimum point during stance phase. | 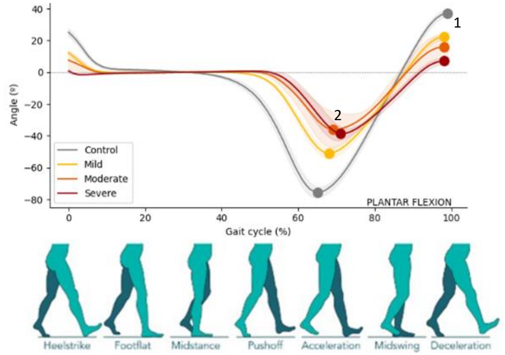  |
| | | **First peak value** (1) (amplitude, time and prominence) | Maximum point of flexion-extension at swing phase. Value and % of cycle when it occurs. | |
| | | First valley value (2) (amplitude, ***time*** (%) and prominence) | Minimum point of flexion-extension at swing phase. Value and % of cycle when it occurs. | |
| | Vertical force (VF) 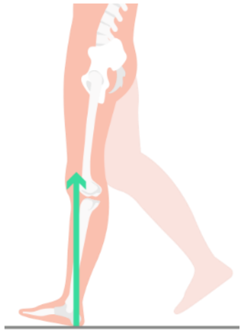 | ***Slope*** (1) | Slope value between maximum and minimum point during stance phase. | 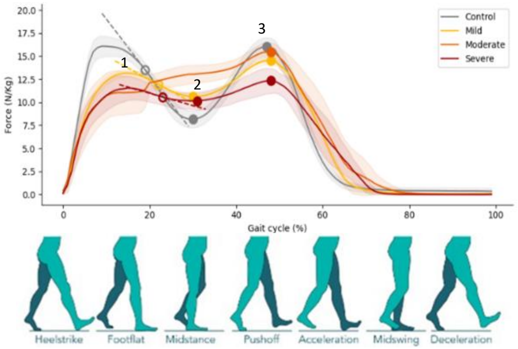  |
| | | **First peak value** (amplitude, **time** (%) and **prominence**) | Maximum point of flexion-extension between heel-strike and middle-stance phase. Value and % of gait cycle when it occurs | |
| | | **First valley value** (2) (amplitude, **time** (%) and ***prominence***) | Minimum point of flexion-extension during heel-strike. Value and % of gait cycle when it occurs. | |
| | | ***Second peak value*** (amplitude, **time** (%) and **prominence**) | Maximum point of flexion-extension between middle-stance and toe-off phase. Value and % of gait cycle when it occurs. | |

| | | | |
|---|---|---|---|
| Centre of pressure (COP) 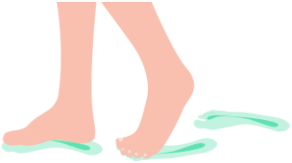 | ***Maximum and minimum value*** | Maximum point on all axis (X,Y). It determines the range of trajectory of center of pressure during gait cycle. | 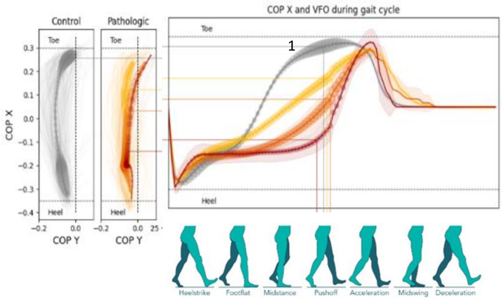  |
| | **Slope between maximum and minimum** | Slope of trajectory of Center of Pressure during gait cycle. | |
| | ***General Center of Pressure slope (1)*** | Slope between the initial moment and heel strike end moment. | |
| | **Loading response Center of Pressure slope** | Slope of 20% of loading response on gait cycle. | |
| | ***Push-off Center of Pressure slope*** | Slope of push-off on gait cycle | |
| All insole signal common features | ***Distance*** and amplitude ***between peaks*** and valleys | Distance in % of cycle between maximums and minimums during gait cycle. Amplitude of the pattern between maximums and minimums during gait cycles. | |
| | **Slope between peaks and valleys** | Slope value between maximum and minimum point during stance phase. | |
| | ***Mean signal*** | Mean value during gait cycle. | |
| | ***Variability signal*** | Standard deviation during gait cycle. | |
| | **Range of motion signal** | Amplitude (maximum-minimum) of the pattern during gait cycle, and per each phase (stance and swing) | |
| | Area of signal | Area under the curve of gait cycle. | |
| Spatiotemporal variables | Distance (m) | Distance covered by the patient during the test. | |
| | Number of steps | Steps walked during the test. | |
| | Speed (m/s) | Velocity of the patient gait. | |
| | Cadence (steps/min) | Number of steps per minute. | |
| | ***Stride length (m)*** | Length in meters of a gait cycle normalized to height. | |
| | ***Stride velocity (m/s)*** | Velocity in meters / second of gait cycle. | |
| | Stance phase (%) | Percentage of time in stance phase, foot-ground contact, on the overall gait cycle. | |
| | Swing phase (%) | Percentage of time in swing phase, foot in the air, on the overall gait cycle. | |
| | Single support (%) | Percentage of time in single support, one foot on the ground, on the overall gait cycle. | |
| | **Double support (%)** | Percentage of time in double support, both feet on the ground, on the overall gait cycle. | |
| | ***Step duration (s)*** | Time token to perform a step. | |

**Supplementary table 6. Definition and representation of variables and features measured by sensors.** Sensor location and explanation of what variable and feature each sensor measures. For this table, it has been represented the curves of angular variables during a gait cycle of ataxia group of patients. Located in the representation curves, significative features obtained for the first objective; in features column in bold, significative features for the correlations study; and in features column on italic, significative features for the longitudinal analysis. Features in bold and simultaneously in italic are those significative in correlations study and in longitudinal study.
